# A behavioral cascade of HIV seroadaptation among US men who have sex with men in the era of PrEP and U=U

**DOI:** 10.1101/2020.11.25.20238758

**Authors:** Steven M. Goodreau, Kevin M. Maloney, Travis H. Sanchez, Martina Morris, Patrick Janulis, Samuel M. Jenness

**Affiliations:** Dept. of Anthropology, University of Washington, Seattle WA; Department of Epidemiology, Rollins School of Public Health, Emory University, Atlanta, GA; Departments of Statistics and Sociology, University of Washington, Seattle WA; Department of Medical Social Sciences, Northwestern University Feinberg School of Medicine, Chicago, IL

**Author notes:** **Corresponding author:** Steven M. Goodreau, Campus Box 353100, University of Washington, Seattle, WA 91895 USA.

**Keywords:** HIV-1, men who have sex with men, seroadaptive behaviors, pre-exposure prophylaxis (PrEP), condom use, treatment as prevention (TasP)

## Abstract

Seroadaptive behaviors help to reduce HIV risk for some men who have sex with men (MSM), and have been well documented in a range of MSM populations. Advancements in biomedical prevention have changed the contexts in which seroadaptive behaviors occur. We thus sought to estimate and compare the prevalence of four stages of the “seroadaptive cascade” in the recent era: knowledge of own serostatus, knowledge of partner serostatus; serosorting (matching by status), and condomless anal intercourse. Serosorting overall appeared to remain common, especially with casual and one-time partners. Although PrEP use did not impact status discussion, it did impact serosorting and the likelihood of having condomless anal intercourse. For respondents not diagnosed with HIV and not on PrEP, condomless anal intercourse occurred in just over half of relationships with partners who were not on treatment. Biomedical prevention has intertwined with rather than supplanted seroadaptive behaviors, while contexts involving neither persist.

## INTRODUCTION

Seroadaptive behaviors represent a longstanding HIV risk reduction strategy practiced by men who have sex with men (MSM) [1-25]. While selection of sexual partners by serostatus (“serosorting”) is most commonly discussed, seroadaptive behaviors include multiple other types of serostatus-based decisions, e.g. selection of sexual acts (e.g. oral vs. anal), selection of sexual role (“seropositioning”, i.e. insertive vs receptive), and decisions around condom use. Akin to HIV care, these can be framed as a “seroadaptive cascade” (**Figure 1**). While seroadaptive behaviors alone are clearly an imperfect strategy for preventing HIV acquisition [26-28], some MSM are more successful in adhering to them than to condom use [29], the latter of which has long been on the decline among MSM overall [15,30]. Two meta-analyses [31,32] found that men practicing serosorting have lower HIV incidence than those who practice condomless anal intercourse (CLAI) without regard to partner status. This confirms that serosorting, and perhaps other seroadaptive behaviors, play an important role towards HIV harm reduction among MSM.

**FIGURE 1:**
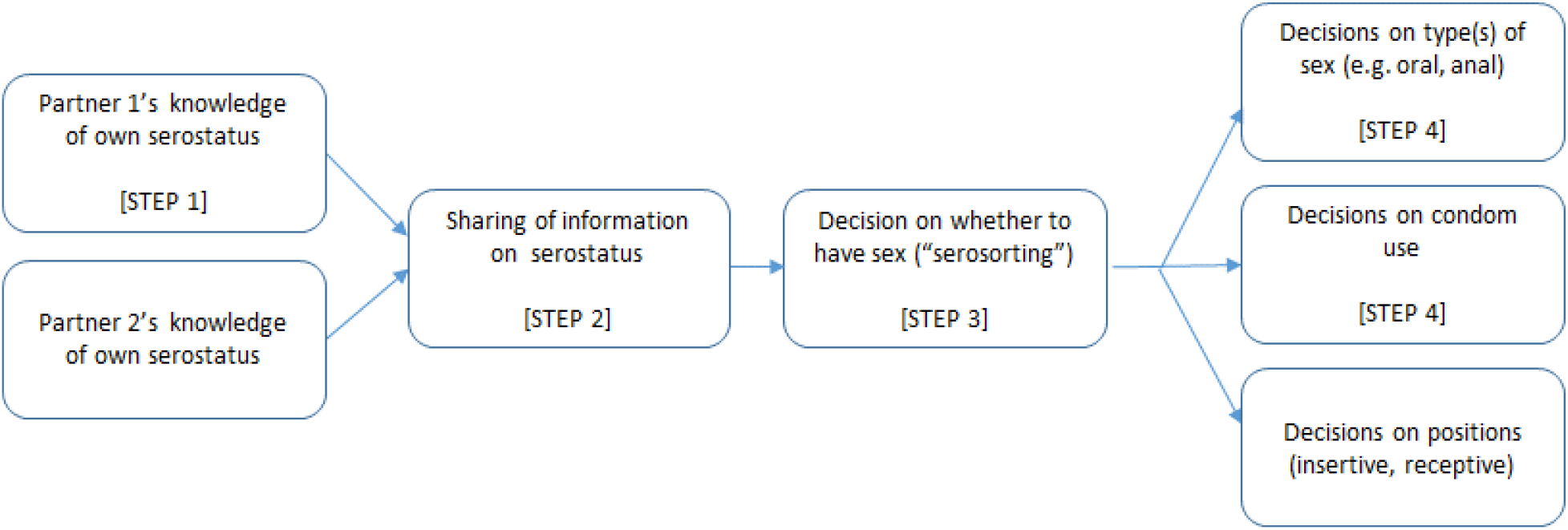
A seroadaptive cascade framework. This provides a schematic of the major elements of seroadaptive behaviors, including the precursor of testing. Steps refer to the components of the analysis in the paper, and do not imply that all of these must occur or, if they do occur, will do so in this exact order. For example, men do not need to know their serostatus to share information (i.e. one can share the fact that one has never tested for HIV), and the decision to have sex can occur without any serostatus discussion at all. In practice, components may also be iterative rather than discretely ordered; for example, a man may change his mind and decide not to have sex if he and a prospective partner cannot agree about sexual type or condom use.

Recent developments in biomedical HIV prevention have fundamentally changed the prevention landscape, and thus the contexts shaping the adoption of seroadaptive behaviors. Both pre-exposure prophylaxis (PrEP) and treatment as prevention (TasP, known commonly as “undetectable = untransmittable”, or “U=U”) work to prevent the spread of HIV through the actions of a single individual. That is, a man taking PrEP as recommended might presume that he has high, consistent protection from HIV acquisition, regardless of his partner’s status. A similar situation holds for a man with HIV who is adherent to ART and has exhibited durable HIV viral load suppression, with respect to transmission. This may lead to men in either group deciding that they need not engage in explicit discussion of HIV status, nor adapt behavior based on perceived status concordance or discordance. We might thus imagine that these biomedical tools have influenced seroadaptive behaviors for these men, and possibly even for men who are HIV-negative and not on PrEP as well through broader changes in safer-sex norms. Seroadaptation appears to have declined overall as PrEP has expanded in San Francisco [33], and subsequent analyses that compared men on PrEP to those off have found some evidence for lower levels of seroadaptive behaviors among the former [34,35].

In practice, not all men on PrEP or ART maintain perfect adherence [36-38] or full self-awareness of their adherence level [39,40], and cases of acquisition by those adherent to PrEP, although rare, do exist [41]. Since individuals may vary in their conceptualizations of risk and the confidence they place in any individual strategy, some proportion may continue to engage in seroadaptive behaviors in conjunction with biomedical strategies. These types of decisions likely play out differently by type of relationships (e.g. main, casual, or one-time) [23], given differences in level of trust around accurate disclosure [42,43], or in trade-offs between prevention and intimacy [44,45]. Given all of the considerations, detailed assessments of current seroadaptive behaviors for MSM across serostatuses or biomedical prevention engagement in multiple settings can provide insight about the current contexts for continued potential HIV transmissions.

Measuring seroadaptive behaviors is complicated by multiple factors. For one, observed behavioral patterns may emerge from other causes besides explicit intentionality [3,19,29,46]. For instance, a preponderance of seroconcordant partners could stem from assortative mixing not on serostatus but on attributes that correlate with it, such as age and race/ethnicity; however, some of the effects on epidemiology may be similar regardless of the actual drivers of partner selection. Another issue is endogenous transmission—i.e. that partners match in HIV status not because of selection but because initially serodiscordant relationships transition to seroconcordant-positive via transmission within the relationship. Finally, knowledge of both self and partner serostatus is generally imperfect and time-varying (until someone is diagnosed with HIV and discloses this). Nevertheless, when considering how knowledge of status impacts behavior, that knowledge itself is a relevant measure, regardless of whether it matches true serostatus.

In this paper, we measure the state of seroadaptive behaviors among MSM during the era of PrEP and U=U. We interrogate four steps in the seroadaptive cascade: HIV testing, disclosure, partner selection, and sexual act/condom use selection. When appropriate, we disaggregate HIV-negative respondents by PrEP status. Our primary hypothesis is that men on PrEP will exhibit less seroadaptation than men who are HIV-negative but not on PrEP. We predict that men who report not knowing their own HIV status will exhibit the lowest levels of seroadaptation. Predicting the relative levels of seroadaptivity between men on PrEP and those with HIV is less straightforward. Regardless, we hypothesize that for all groups, both knowledge of and concordance in serostatus will be highest in more long-term relationships. By considering each stage and the relational types in which they occur, we aim for our work to provide the evidence needed to parametrize further quantitative analyses, such as mathematical modeling or the development of individual or relational risk scores, that can help to assess and thus intervene upon the contexts in which probable transmission events continue to occur in the biomedical prevention era.

## METHODS

We used data from the ARTnet study, a web-based sexual network survey of MSM conducted in conjunction with the long-standing American Men’s Internet Survey (AMIS) in two waves (Jul. 2017–Feb. 2018 and Sep. 2018–Jan. 2019). Detailed methods are published for both ARTnet [47] and AMIS [48-51]. Recruitment for AMIS occurred through banner ads on websites and social network applications popular with MSM. Respondents who completed AMIS were requested to participate in ARTnet, which included additional questions on sexual networks and behaviors with male partners. Eligibility criteria for ARTnet included being aged 15–65, assigned male sex at birth, having cisgender male identity, and having any lifetime sexual activity with another man.

Most of our outcomes of interest used relationships as the unit of analysis. Respondents were asked summary questions about their relationship history and detailed questions about their most recent male partners (up to 5) within the last year. Relationships were categorized into three types: main (“someone that you feel committed to above all others, someone you might call your boyfriend, significant other, life partner or husband”), casual (anyone else a respondent has had sex with more than once); and one-time.

Respondents were categorized by self-reported knowledge of HIV status: *HIV-positive* (those who had ever had a positive HIV test); *HIV-negative* (those who had tested but never with a positive result); and *HIV-unknown* (those who had never tested, never received their results, or reported not knowing their results). Partners were similarly categorized by the respondent’s reported knowledge of their serostatus; here, HIV-unknown included cases where respondent said their partner had never been tested, their partner did not know his status, or that they did not know their partner’s status.

Respondent’s PrEP use was categorized on a per-partnership basis, i.e. whether they were taking PrEP during none, some, or all of the relationship. For behaviors by PrEP use, we compared cases with PrEP use during all or none of the relationship, since with partial PrEP use we cannot tell which behaviors occurred during the period with and without PrEP. To determine the impact of this decision, we repeated the relevant analysis as any PrEP versus no PrEP during the relationship (see online Supplement). Partner PrEP and ART use were measured similarly, with our analyses also comparing use during all or none of the relationship.

We considered four steps of the seroadaptive cascade (**Figure 1**): knowledge of own status (Step 1), knowledge of partner’s status (Step 2), serosorting (Step 3), and selection of sexual acts (CLAI vs others; Step 4). We did not address seropositioning, as previous work found that this was much less common than other seroadaptive behaviors using either behavioral or intentionality definitions, and also had low consistency between those definitions [46].

At all cascade steps, we present our results in terms of the prevalence of each examined behavior by sub-group. This satisfies one of our goals: to provide a rich set of parameters for those modeling bio-behavioral prevention strategies among MSM in the presence of both PrEP and viral suppression. We then present tests of our hypotheses using exact binomial tests to calculate 95% confidence intervals on estimates with a binary outcome, and Fisher’s exact tests to compare estimates across groups.

## RESULTS

Our inclusion criteria from the broader ARTnet study yielded 4,512 respondents, reporting on 13,800 relationships, or 3.1 relationships per respondent. These were slightly reduced relative to previous published analyses, since we excluded 392 respondents not asked about PrEP use early in the first wave. **Table 1** lists descriptive statistics for the samples of both respondents and relationships. Overall, 9.5% reported being diagnosed with HIV. One-time contacts represented just over half of reported relationships; main and casual relationships, with their longer durations, still reflected most of the relationship time and sexual acts. There were large numbers of relationships in which the respondent reported being HIV-negative and on PrEP (n=2,239) or HIV-negative and not on PrEP (n=7,937) throughout the relationship. However, the relationships reported by men diagnosed with HIV mostly comprised cases where they were on ART throughout (n=1,126) with very few cases of no ART use (n=69). We thus did not disaggregate any analyses of behaviors of HIV-positive respondents by own suppression status.

**TABLE 1:** Descriptive statistics for respondents and relationships

|  | Individual-level<br>(n = 4,512) | Relationship-level<br>(n = 13,800) |
| --- | --- | --- |
| Respondent median age [IQR] | 34 [24-49] | 33 [24-49] |
| Race/ethnicity of respondent |  |  |
| Black (non-Hispanic) | 232/4512 ( 5.1%) | 615/13800 ( 4.5%) |
| Hispanic | 626/4512 (13.9%) | 2005/13800 (14.5%) |
| White (non-Hispanic) | 3250/4512 (72.0%) | 9988/13800 (72.4%) |
| Others | 404/4512 ( 9.0%) | 1192/13800 ( 8.6%) |
| Self-reported HIV status of respondent |  |  |
| HIV-negative | 3334/4512 (73.9%) | 10680/13800 (77.4%) |
| HIV-positive | 428/4512 ( 9.5%) | 1300/13800 ( 9.4%) |
| Don't know | 750/4512 (16.6%) | 1820/13800 (13.2%) |
| Relationship type |  |  |
| Main |  | 2076/13800 (15.0%) |
| Casual |  | 4751/13800 (34.4%) |
| One-time |  | 6973/13800 (50.5%) |
| Ever taken PrEP (HIV-neg. respondents) |  |  |
| Yes |  |  |
| No | 817/3334 (24.5%) |  |
|  | 2517/3334 (75.5%) |  |
| On PrEP during this relationship (HIV-neg. respondents) |  |  |
| Entire time |  | 2239/10680 (21.0%) |
| Some of the time |  | 448/10680 ( 4.2%) |
| None of the time |  | 7937/10680 (74.3%) |
| Not sure/no answer |  | 56/10680 ( 0.5%) |
| On antiretroviral medication during this relationship (HIV-pos. respondents) |  |  |
| Entire time |  | 1126/1300 (86.6%) |
| Some of the time |  | 48/1300 ( 3.7%) |
| None of the time |  | 69/1300 ( 5.3%) |
| Not sure/no answer |  | 57/1300 ( 4.4%) |

IQR = interquartile range

### Step 1

**Figure 2** shows multiple measures for respondents’ awareness of their own status. The proportion of respondents who reported ever having an HIV test rose rapidly from less than one in five in the mid-teens to near-universality around age 30, at which point it asymptoted with strong consistency across ages. Including all ages, the proportion with a test ever was 84.0%, but including only respondents 25+ raises this to 92.9%, or for 30+ to 94.4%. Testing in the last two years followed the same pattern at young ages—when nearly all testing would be recent—but diverged in the mid-20s, and experienced absolute decline beginning around age 30. This trend was presumably driven in part by the increasing number of men who have tested positive by age, and thus do not require additional testing; this sub-group rose steadily with age until about 50, above which 19.9% of respondents reported having an HIV diagnosis. To determine the extent of this effect, we also plotted the proportion of men having a test in the last 2 years only among those who had never tested HIV-positive. This proportion suggests three rough phases in testing across the lifecourse for MSM who remain HIV-negative: one up through the mid-20s when testing is ramping up; one from then until near age 50 when testing is roughly stable, and one from age 50 when testing is in decline. Within the middle group, 82.7% had tested in the last two years. Collectively, these findings emphasize that, in subsequent analyses, respondents of “unknown” status largely reflect younger MSM, while those of positive status disproportionately reflect older men.

**FIGURE 2:**
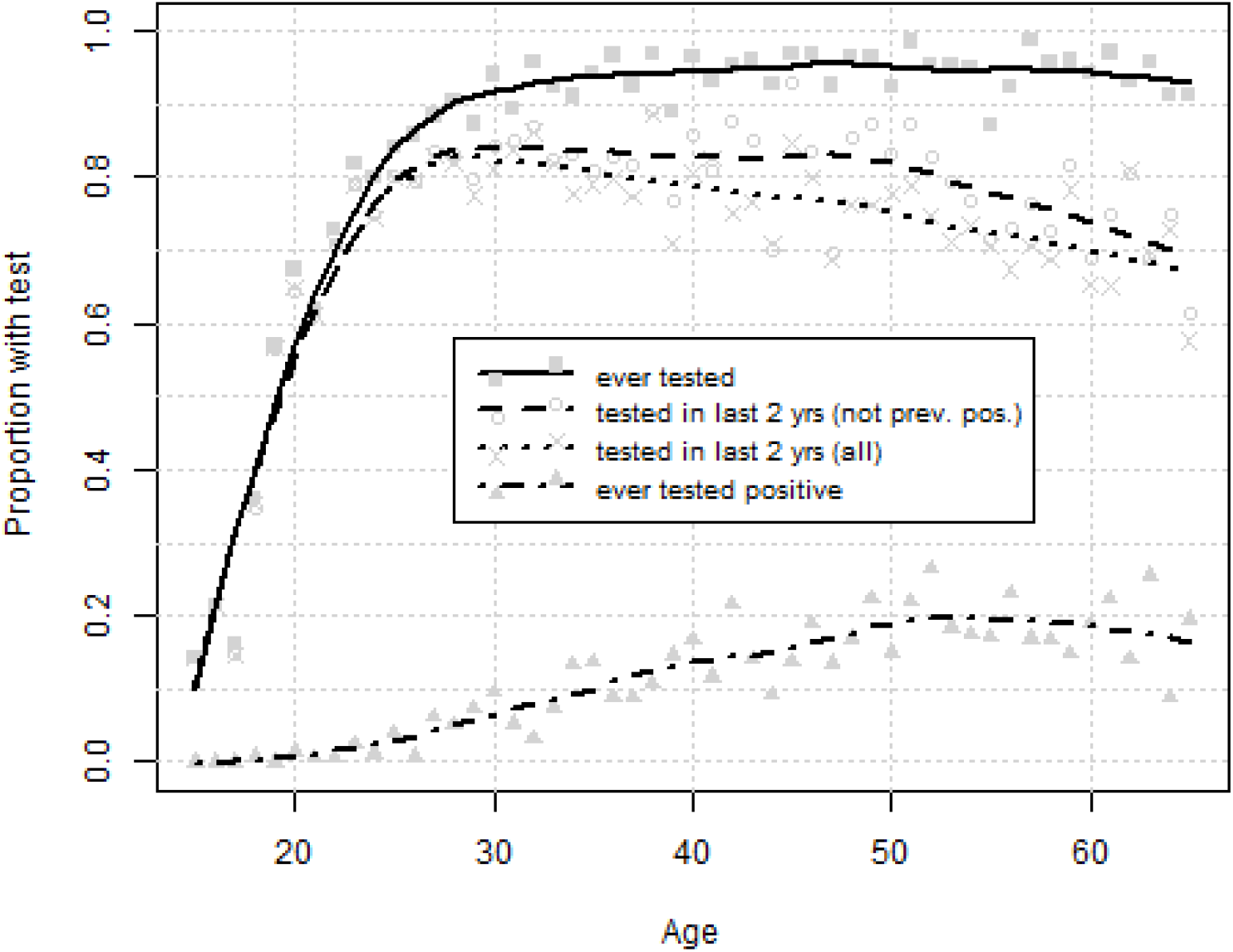
Measures of respondents’ knowledge of their own HIV status, by age. From top to bottom, metrics reflect the proportion at each age who report ever having had an HIV test; the proportion who have had an HIV test in the last 2 years, out of those who have not previously been diagnosed positive; the proportion overall who have had an HIV test in the last 2 years; and the proportion who have ever tested positive for HIV. Dots reflect the mean value for respondents of a given age. Lines represent loess curves with α=0.5, as implemented with the *loess* command in R v. 4.0.2.

### Step 2

**Figure 3a** depicts the proportion of partners for whom the respondent claims knowledge of their partner’s serostatus, regardless of what that status was. Results are divided by respondent serostatus, PrEP status for negative respondents, and relational type. Overall, respondents reported knowing partner status in 68.3% of relationships. As hypothesized, the likelihood of knowing a partner’s status declined with less committed relationship types, regardless of respondents’ serostatus or PrEP status (Fisher’s exact test, p <1e-7 in all cases). We further hypothesized that negative men on PrEP would report lower status communication than negative men not on PrEP for a given relational type, especially for non-main relationships; however, we did not find evidence for a significance difference between these two populations (one-tailed Fisher’s exact test, p>0.75 for each relational type). Men on PrEP were more likely to know partner status than HIV-positive men were for casual and one-time partners, but not for main partners. Men who did not know their own HIV status reported the least knowledge of partner status overall; however, we note that even these men reported knowing their one-time partners’ status fully half of the time. Redoing this comparison between relationships during which the respondent was never on PrEP versus ever (as opposed to always) on PrEP yielded qualitatively similar findings (Figure S1).

**FIGURE 3:**
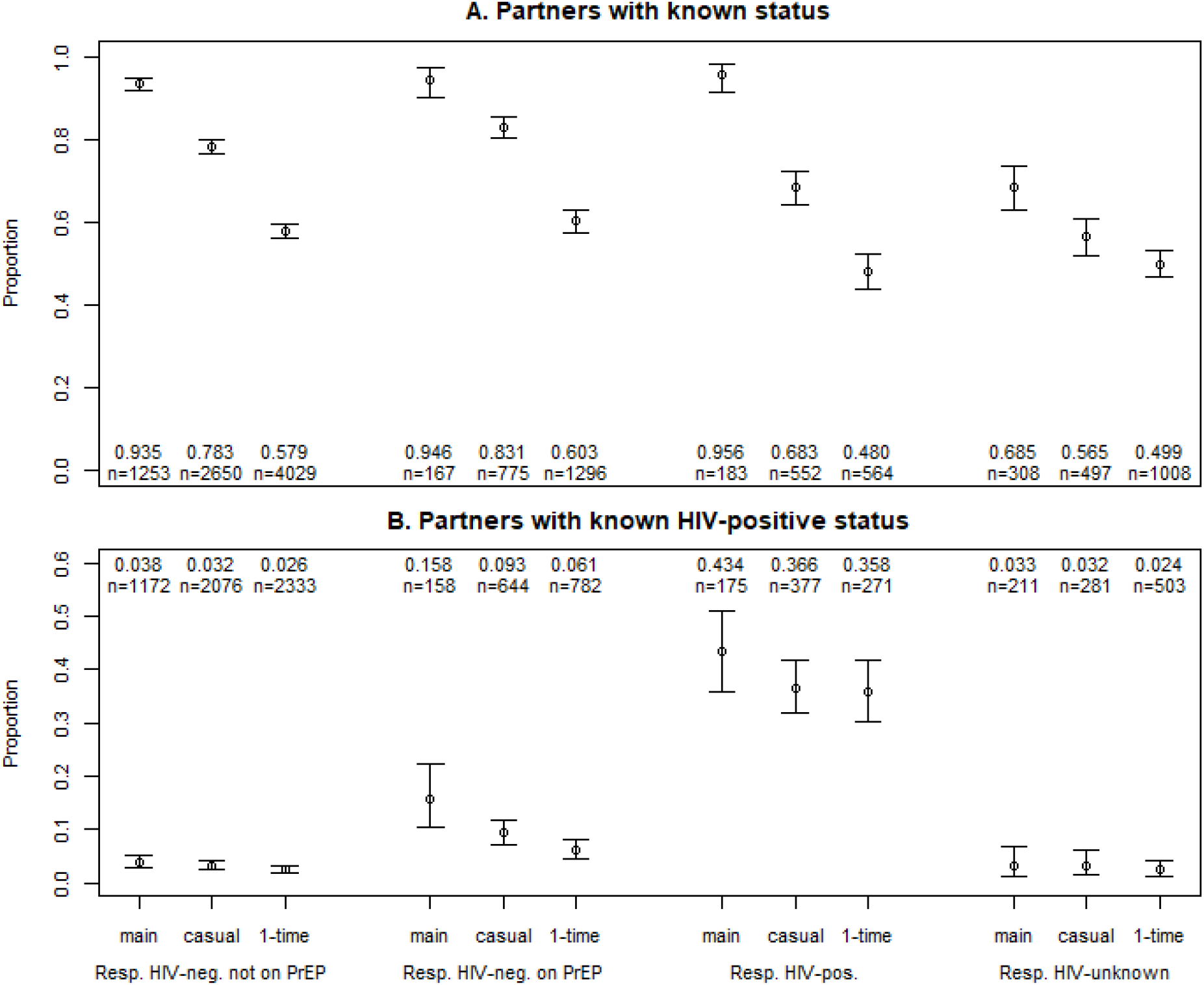
Partner status, by respondent HIV status and relationship type: (a) Proportion of partners whose HIV status respondents report knowing; (b) Proportion of partners who respondents report as HIV-positive, among partners of known status. CIs = binomial proportion confidence intervals, selected due to the binary outcome and the small sample sizes in some categories. Relationship types are defined in the text. Note the difference in scale on the *y*-axis to make visible the differences by relational type in Figure 3B.

### Step 3

The largest distinction in partner serostatus (conditional on the respondent reporting it as known) lay in the considerably higher proportion of HIV-positive partners for HIV-positive respondents over any other type of respondent, for all three partner types (**Figure 3b**), consistent with serosorting. As predicted, HIV-negative men on PrEP reported a higher proportion of known HIV-positive partners than did negative men not on PrEP (RR = 2.7 overall). This association between respondent PrEP status and partner HIV status was particularly strong for main partners, with a RR of 4.2, compared to 2.9 for casual and 2.3 for one-time; however, all three were significant relative to the null of no difference by PrEP status (Fisher one-tailed exact tests, p<1e-5 for all relationship types). Men on PrEP were the only population to show a significant difference in their proportion of HIV-positive partners across partner type (Fisher’s exact test, p=3e-4). Those who reported not knowing their HIV status had responses most similar to HIV-negative men not on PrEP. Again, reanalysis to include PrEP use at any point in the relationship did not change the qualitative results (Figure S1).

### Step 4

For act type and condom use behaviors, we focused on disaggregating by partner status rather than partner type, as the former is fundamental to the transmission potential associated with each behavior (**Figure 4**). HIV-negative respondents on PrEP had higher proportions of relationships with any CLAI than did HIV-negative respondents not on PrEP, for all partner serostatuses (one-tailed Fisher’s exact test, p<1e-6 in all cases). Indeed, the numbers for respondents on PrEP were similar to those of HIV-positive men (p>0.30 in all cases). Again, responses for unknown-status respondents were most similar to those for HIV-negative men not on PrEP.

**FIGURE 4:**
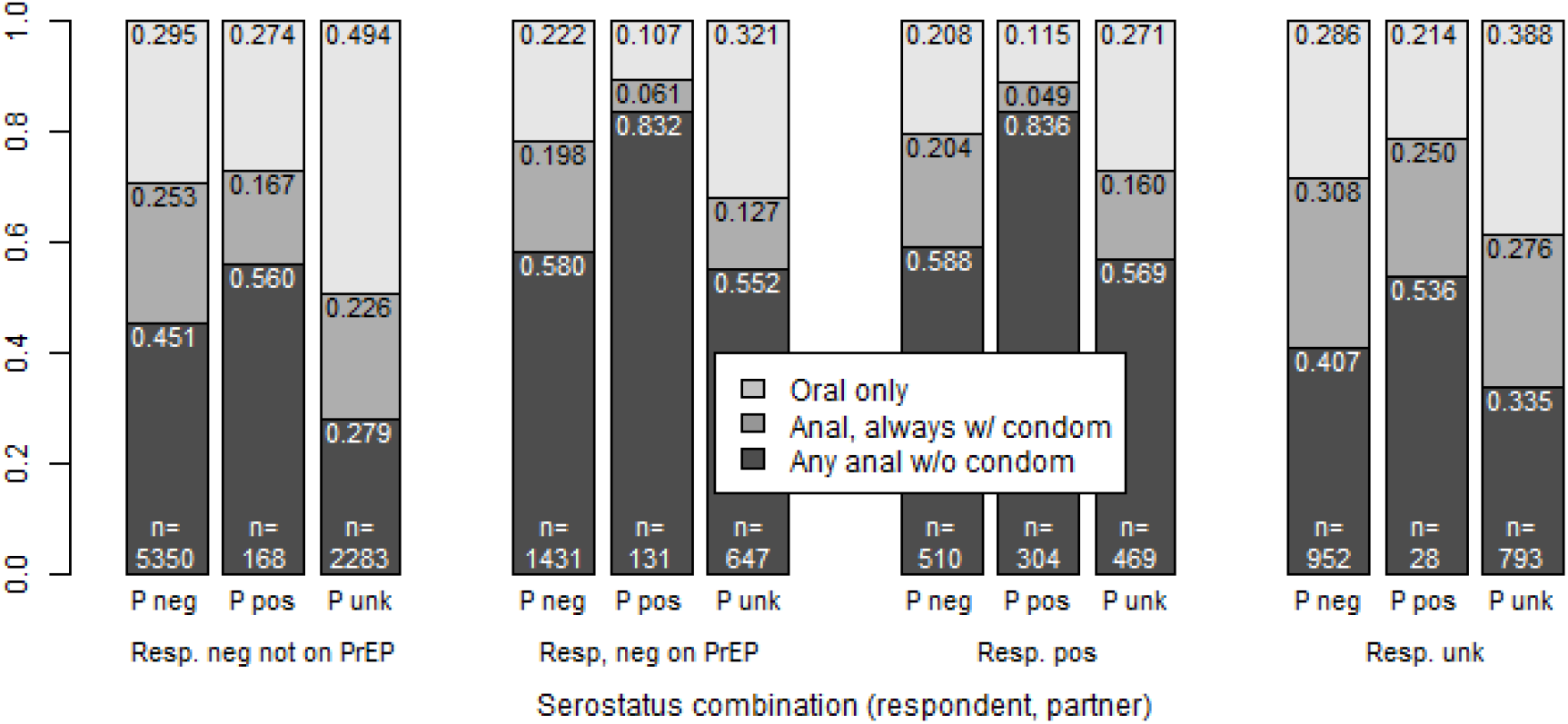
Sexual acts and condom use within relationships, by respondent HIV status and PrEP use and partner HIV status. Relationships are categorized by act with highest transmission probability, i.e. a relationship listed as “anal, always w/ condom” may also contain oral sex acts, while “any anal without condom” may also contain anal sex with condom and/or oral sex. P neg = partner HIV-negative; P pos = partner HIV-positive; P unk = partner HIV status unknown

We note a consistent trend that we did not predict; for all types of respondents, regardless of status or PrEP use, CLAI was most likely to occur with known HIV-positive partners. Of course, the implications for this differ for HIV-positive versus HIV-negative men on or off PrEP. More than half of the relationships between HIV-negative respondents not on PrEP and their HIV-positive partners involved some CLAI, and the same was true for serostatus-unknown respondents. Since these types of relationships may entail the greatest transmission opportunity, we further disaggregated them by the respondent’s report of their HIV-positive partner’s use of ART. We did not see a significant difference among HIV-negative respondents not on PrEP in the probability of CLAI with HIV-positive partners on ART (56.5%, 95% CI = 47.0%–65.7%, n=115) versus off ART (53.1%, 95% CI = 38.3%–67.5%, n=49). We considered the same metrics for respondents on PrEP and also saw no significant difference (partners on ART: 80.0%, 95% CI = 71.3%–87.0%, n=110; partners off ART, 90.0%, 95% CI = 68.3%–98.8%, n=20). One potential reason for the relatively high likelihood of CLAI with HIV-positive partners by all respondent types might be that they are more likely to be main partners than those of HIV-negative or HIV-unknown status, as seen in Figure 3. Table S1 (online supplement) reveals that, for each type of respondent (HIV-negative on and off PrEP, HIV-positive), CLAI is more likely with an HIV-positive main partner than HIV-positive one-time partner; however, the values for HIV-positive casual partners are similar those for main partners for each group of respondents.

## DISCUSSION

In this study, we investigated four steps of the seroadaptive cascade in the era of biomedical prevention, comparing behaviors variously by serostatus knowledge, biomedical prevention use, and partnership type. We found that PrEP use does not have a discernible impact on status discussion, but does impact partner selection and the likelihood of having condomless anal intercourse. All groups of respondents were most likely to have CLAI with partners diagnosed with HIV, especially with main or casual partners. For respondents not diagnosed with HIV and not on PrEP, CLAI occurred in just over half of relationships with partners who were not on treatment. The probability that these respondents would have CLAI with one of their partners of unknown status was lower, but such partners were also much more common (n=2283, or 17.5% of all relationships). This suggests the persistence of sexual activity within known serodiscordant relationships in which men are aware of the absence of any form of prevention of HIV transmission, either biomedical or behavioral, as well as within many relationships where this absence is a reasonable possibility. Both types represent key areas where identification and intervention continue to be needed, and could have substantial impact.

Our primary finding is broadly consistent with two recent studies that also disaggregated disclosure and partner status by respondent PrEP use. In an analysis that combined national and New York City samples, Grov et al. [34] found that HIV-negative men on PrEP had a higher proportion of positive casual partners than those off PrEP did (roughly 17% and 11% of their partners of known status, estimated from numbers in their Table 3.) Wang et al. [35] found similar numbers (17.1% vs 9.3% of all partners with known status, respectively) in a Montreal-based sample. While the pattern of less serosorting by men on PrEP was consistent across all studies, our numbers suggest overall higher serosorting by HIV-negative men than these previous studies did (8.7% and 3.1% known positive partners for men on or off PrEP in this study when averaged across partnership types, or 7.9% and 2.9% when excluding main partnerships for comparability to Grov et al.) Our sample is younger and more White than the US/NYC study, and more geographically diverse and less urban than the Montreal study, all of which may make HIV prevalence in the partner pool of our respondents lower overall. Because the previous studies did not disaggregate by relationship type, we cannot compare our findings to them in this regard. Our observation that men on PrEP have high rates of main partners known to be diagnosed with HIV (15.8%) relative to other partner types undoubtedly reflects at least some reverse causality—i.e. an HIV-negative man with a main partner living with HIV is indicated for PrEP if they have anal sex, and would have high motivation to take it. For one-time partners, the number is smaller (6.1%) and more similar to but still significantly above that of HIV-negative men off PrEP (2.6%). All of these numbers are far below the comparable numbers for respondents diagnosed with HIV, or indeed the proportion of MSM diagnosed with HIV in the US population [52]. Whether they reflect explicit serosorting by men on PrEP or partner selection on other attributes that correlate with status, the pattern does present evidence for a sexual network that remains fairly segregated between men who are or are not diagnosed with HIV, even as biomedical prevention has been touted as an option to allow individual MSM to more comfortably “bridge the serodivide” [53].

In contrast to our findings, these same two studies found that men with recent PrEP use were significantly *more* likely to know their partner’s status than HIV-negative men without recent PrEP use (83% versus 74% for Grov et al.; 69.4% versus 50.5% for Wang et al.) Our comparable numbers, averaging across partner types included by each study, were 71.1% and 66.0% (for Grov et al.) and 74.0% and 70.4% (for Wang et al.) These trended in the same direction but were closer and not significantly different from each other. Regardless, all three studies agree that men on PrEP are not generally considering the protection it affords as a reason to engage in *less* serodiscussion. While we confirmed the expected trend by relational type, we also found that men on PreP were more likely than not (60.3)% to report knowing even their one-time partners’ HIV status. While one might imagine that knowledge of a main partner’s serostatus would reflect overall intimacy and not solely concern for HIV transmission risk, this should be less true for casual and most especially one-time partners. The high rates of reported status knowledge by men on PrEP here may reflect multiple phenomena: (1) a continued desire by men on PrEP to assess their HIV risk, perhaps because of concern about their own adherence or other PrEP failures; (2) disclosure in the form of dating app profiles, which would occur automatically and without need for explicit discussion [54]; and (3) the mutuality of HIV status disclosure, such that men on PrEP are sharing and receiving HIV and biomedical status information for their partner’s benefit [55]. The first of these includes cases where men on PrEP are not only seeking partners who are HIV-negative, but who themselves are on PrEP, to provide yet another layer of protection (“PrEP-sorting”); while we did not investigate this pattern here, both Grov et al. and Wang et al. found evidence for this phenomenon. Evidence for the second and third possibilities may come from our observation that men who report not knowing their own status still reported knowing their one-time partners’ status more than half the time. It also highlights the possibility that some men report their status as negative to potential partners as long as they have never had an HIV-positive test result—even if they have never tested or tested very long ago [56].

We found that the first precursor for seroadaptation—having an HIV test—remains common, as one might expect given that accessing new biomedical prevention modalities still requires HIV screening, and the continued emphasis on testing by public health campaigns targeted at MSM. For MSM aged 25–49 who have not been diagnosed with HIV, roughly 5 out of 6 have tested in the last 2 years. However, those below age 25 show considerable room for improvements in testing, with only 58.4% aged 15–24 testing in the last 2 years, or 63.8% of those 18–24. This latter number is well below the 78.8% of MSM aged 18–24 who had tested in the last 12 months in the most recent NHBS results [52], despite the latter testing time window being shorter. This may reflect differences in web-based versus venue sampling, and requires further investigation.

We found that men on PrEP were just as likely to have CLAI with their partners diagnosed with HIV (83.2%) as were respondents who themselves were diagnosed with HIV (83.6%). Since men on PrEP appeared to exhibit some level of seroadaptivity in terms of partner selection, we might then also anticipate that they would do so in terms of act selection and condom use; the high probability here and its similarity to the rate for respondents diagnosed with HIV suggests this does not appear to be the case.

Respondents of all serostatus and biomedical prevention status reported that CLAI was most likely with their partners who had diagnosed HIV. This finding for HIV-negative respondents implies that something other than simple seroadaptive behaviors is occurring. One likely explanation is selectivity—where those with the strongest propensity for CLAI acquire HIV at disproportionate rates and continue that propensity with their partners, including HIV-negative ones. Nevertheless, it is notable that this apparent effect is strong enough to reverse any tendency for selective use of condoms by HIV-negative men not on PrEP with their partners diagnosed with HIV.

## Limitations

Our study relies on a convenience sample, as do all national surveys of MSM to some extent. Non-Hispanic White men were slightly over-represented, relative to the adult US male population, as were younger men. Nevertheless, a previous analysis of this data set found that, after accounting for unknown responses, HIV prevalence in the sample was in line with national estimates [47], and our median age matched that in the previous similar studies to which we compared our results [34,35]. It remains challenging to know how different respondents may interpret questions such as knowledge of partners’ status—e.g., where some may assume a one-time partner’s HIV-negative disclosure as truth while others report it as unknown. Again, however, it is presumably respondents’ own perception of partner’s status that most directly influences potential seroadaptive behavior. Answers to our questions were not necessarily the same way respondents would describe their status to a partner, especially for those whose last negative test was long ago. As with most serosorting studies, we do not have information on partnerships that did *not* occur specifically because of serodiscordance. Our inclusion of multiple hypothesis tests increases the overall Type 1 error rate and may yield false positives; we note, however, that all significant differences had *p*-values orders of magnitude below our significance level, and would thus hold up under a multiple comparison correction.

## Conclusions

This study provides substantial new information on the recent magnitude of testing, disclosure, serosorting, sexual act selection, and use of condoms among US MSM. Our future work will incorporate this information into mechanistic transmission models, along with measures of imperfect adherence to PrEP and ART, to obtain estimates of the attributable fraction of transmissions among MSM occurring in different contexts, i.e. by relationship type and by the biomedical prevention methods used by the men in these relationships. Such work would be relevant for HIV as well as for other major reportable STIs (e.g., syphilis), which circulate on the same sexual network as HIV. With more than 25,000 new HIV diagnoses among US MSM each year still, such models are critical to identify the conditions where prevention efforts—biomedical and behavioral—remain insufficient, as we work towards the goal of ending the HIV epidemic.

## Data Availability

Data are available for use by researchers at https://github.com/EpiModel/ARTnetData. Data access requires signing a Memorandum of Understanding given the sensitive nature of some survey questions.

## ACKNOWLEDGMENTS

The authors would like to thank the study participants, the full AMIS and ARTnet research teams, and the Network Modeling Group at the University of Washington.

## COMPLIANCE WITH ETHICAL STANDARDS

### Research involving Human Participants

This study was approved by the Emory University Institutional Review Board. It was performed in accordance with the ethical standards laid down in the 1964 Declaration of Helsinki and its later amendments.

### Informed consent

Informed consent was obtained from all individual participants included in the study.

