## Supplement for "A behavioral cascade of HIV seroadaptation among US men who have sex with men in the era of PrEP and U=U"

**Supplementary material**

We initially conducted Steps 2 and 3 of our analyses by excluding cases where respondents were on PrEP for only part of a relationship, as it was impossible to determine whether any specific types of sex acts within that relationship (e.g. condomless anal intercourse) occurred exclusively while on PrEP, exclusively while off PrEP, or both. However, the relationships excluded by this decision are biased towards longer relationships, especially those extending back in time before the widespread availability of PrEP. To determine the extent of the impact of this decision on our overall findings, we repeated the analysis by combining the relationships with partial PrEP use with cases of full PrEP use (i.e. distinguishing between any PrEP use versus no PrEP use). Figure S1 below displays these results; not that only the section for respondents on PrEP differs from Figure 3 in the main text.

Sample size increases by 97% for main relationships (329 versus 167), 33% for casual relationships (1032 versus 775), and by 2% for one-time relationships (1,325 versus 1,296). The very small number of new cases in the last set reflect some form of respondent error, as one-time relationships by definition involve either PrEP use or non-use throughout their duration.

Patterns in our outcomes of interest are overall qualitatively simular to Figure 3; point estimates in panel A (proportion of partners with known status) are 0.951, 0.844 and 0.607 by partner type here, compared to 0.946, 0.831 and 0.603 in Figure 3 panel A. Comparable numbers for panel B (proportion of partners known to have HIV) are 0.128, 0.093 and 0.063 here and 0.158, 0.093 and 0.061 in Figure 3. The significance or non-significance of all relevant statistical tests matches that performed on the reduced sample in the main text.

**Figure S1**

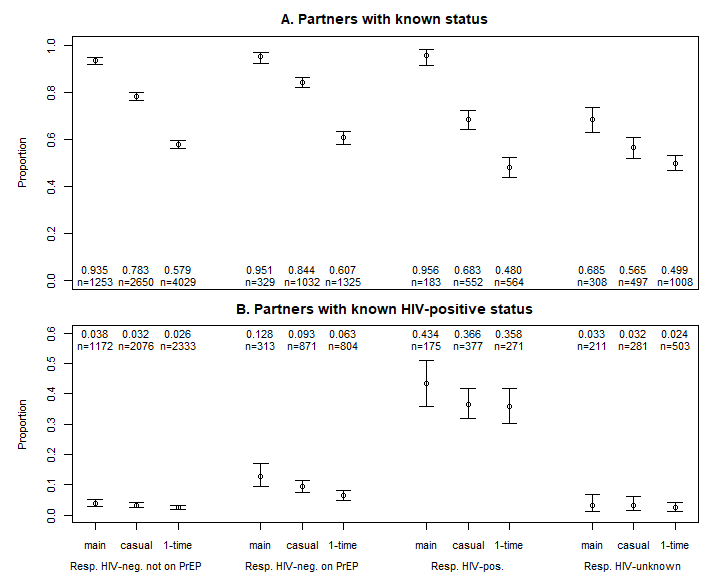

Table S1: CLAI with known HIV positive partners, by respondent self-reported serostatus and relational type

| Resp. serostatus | Main partners | Casual partners | One-time partners |
| --- | --- | --- | --- |
| HIV-negative no PrEP | 65.9% (n=44) | 65.2% (n= 66) | 37.9% (n=58) |
| HIV-negative PrEP | 96.0% (n=25) | 91.7% (n= 60) | 65.2% (n=46) |
| HIV-positive | 86.1% (n=72) | 86.0% (n=136) | 78.1% (n=96) |

We exclude HIV-unknown respondents, as their total number of partners across the three categories (28) is too low to calculate meaningful proportions in each one
